# Coronary perforation occurring during percutaneous coronary intervention is associated with persistent high inpatient mortality and complications

**DOI:** 10.1101/2024.01.04.24300874

**Authors:** Mohammad Reza Movahed, Nishant Satapathy, Mehrtash Hashemzadeh

## Abstract

**Background:** Coronary perforation is one of the major complications of percutaneous coronary intervention (PCI). The goal of this study was to evaluate adverse outcomes and mortality in patients suffering from coronary perforation during PCI above the age of 30.

**Methods:** The National Inpatient Sample (NIS) database, years 2016-2020, was studied using ICD 10 codes. Patients suffering from perforation were compared to patients without perforation during PCI.

**Results:** PCI was performed in a weighted total of 10,059,269 patients. Coronary perforation occurred in 11,725 (0.12 %) of all PCI performed. The mortality rate of patients with perforations was very high in comparison to patients without perforations. (12.9% vs 2.5%, OR: 5.6, CI:5-6.3 p<0.001). Furthermore, patients with coronary perforations had much higher rates of urgent coronary bypass surgery, tamponade, cardiac arrest, and major cardiovascular outcomes. Mortality remained high and over 10% in the 5-year study period.

**Conclusion:** Using a large national inpatient database, all-cause inpatient mortality in patients with coronary perforation is very high (over 10%) with persistently high mortality rates over the years study suggesting that treatment of perforations needs further improvement.

## Introduction

Percutaneous coronary intervention (PCI) is predominant a nonsurgical treatment option for coronary artery disease with appropriate indication (CAD)^1^. PCI is a major indication in patients with acute ST-elevation myocardial infarction (STEMI) and Non-ST-elevation acute coronary syndrome (NSTE-ACS)^1,2,4^. As of 2017, there are more than 230 PCIs per 100,000 people in the US^3^. Despite its widespread use, PCI is less indicated in stable CAD and is mostly done for symptom relief. PCI is associated with cardiac and vascular complications, one of which is coronary perforation is a major one^7^. Defined as a tear in the coronary artery during PCI, coronary perforations are classified as one of three classes. The first, an “extraluminal crater without extraversion”, and second, “pericardial or myocardial blushing,” can be treated conservatively, while the third class, a perforation with a diameter greater than 1 mm, has been deemed the most life-threatening^12,13^. Despite having an incidence rate of 0.5%, coronary perforation continues to be a major complication of PCI and has been associated with an increased incidence of MI, CABG, and mortality^8,9,10^. In addition, individuals who experienced perforation were at an increased risk for complications prior to the procedure, such as MI, cardiogenic shock, stroke, and pericardial tamponade^9^. 1 in 250 PCIs still lead to coronary perforations^6^. There remains a need for a large database analysis of clinical outcomes and mortality rates for individuals experiencing perforations due to PCI^9,10-17^. Using the National Inpatient Sample (NIS), this study evaluates adverse outcomes and mortality in patients suffering from coronary perforation during PCI above the age of 30.

## Methods

### Data Source

Deidentified, publicly available patient data was obtained from the National Inpatient Sample (NIS), Healthcare Cost and Utilization Project (HCUP), and Agency for Healthcare Research and Quality. The dataset includes discharge data for 2,011,854 patients (10,059,269 after discharge weights were applied), which counts to a sample of 20% of the discharges from U.S. hospitals and close to 98% of the total U.S. population. Due to the open-source nature of the data without any patient identifier, our study is exempt from institutional review.

### Study Population

Patient data from 2016-2020 was analyzed for this study. International Classification of Diseases, Tenth Revision, Clinical Modification (ICD-10-CM) and International Classification of Diseases, Tenth Revision, Procedure Coding System (ICD-10-PCS) codes were used to identify patients who had PCI. These ICD-10-PCS codes were 02703(4-7)Z, 02703(D-G)Z, 02703TZ, 02713(4-7)Z, 02713(D-G)Z, 02713TZ, 02723(4-7)Z, 02723(D-G)Z, 02723TZ, 02733(4-7)Z, 02733(D-G)Z, 02733TZ, 02H(0-3)3DZ, 02H(0-3)3YZ, 027(0-3)3ZZ, 02C(0-3)3Z7, 02C(0-3)3ZZ, 02F(0-3)3ZZ. The ICD-10-CM code I97.51 was then used to identify patients who had the adverse event of coronary perforation. All patients analyzed had an age above 30 years. Age, gender, and race were then used to for demographic analyses.

### Codes for study Outcomes

Outcomes analyzed included mortality and complications, specifically pericardial effusion (I31.3), cardiac tamponade (I31.4), acute lactic acidosis (E87.20), sepsis (A40, A41, R65.2, T81.4, T83.7, A48.3, B37.7, A54.86), nontraumatic ischemic infarction of muscle (M62.2), atherosclerosis of extremity arteries with rest pain (I70.22), atherosclerosis of extremity arteries with gangrene (I70.26), ischemia/infarction of the kidney (N28.0), injury of the kidney (S37.0), acute kidney failure (N17.9), postprocedural kidney failure (N99.0), acute posthemorrhagic anemia/anemia following acute blood loss (D62), elevation of lactic acid dehydrogenase (R74.02), hemorrhage, not elsewhere classified (R58), acute postprocedural respiratory failure (J95.821), disseminated intravascular coagulation (D65), mechanical complications of other cardiac and vascular devices and implants (T82.5), blood transfusion (30233N, 30243N, 30233P, 30243P, 30250P, 30260P, 30250N, 30260N, 30253P, 30263P, 30253N, 30263N, 30230N, 30240N, 30230P, 30240P), cardiac arrest (I46, I97.71, I97.12), and stroke (I60-I63, I65, I66).

### Statistical Analysis

Patient demographics, specifically age, gender, race, and median income, clinical and hospital characteristics were analyzed as means and 95% confidence intervals were provided. Chi-squared analyses provided results to analyze trends over time for categorical variables. To analyze trends over time for continuous variables, univariate linear regression was used. Multivariable logistic regression yielded values for clinical outcomes relative to patient and hospital characteristics and their odds over time. Significance was determined as 2-sided p-values where p<.05 was defined as statistically significant. STATA 17 (Stata Corporation, College Station, TX) was used for all statistical analyses.

## Results

A total of 10,059,269 patients underwent PCI, of which 11,725 (0.12%) experienced a coronary perforation. The average age of the subjects in the study was 70.04 (CI: 69.98 – 70.04), where individuals aged 61+ made up a majority of the cohort (77.84%). 63.68% of the cohort were men and 36.32% were women. Moreover, 77.03% of the study cohort was Caucasian. (*Table 1*). The most common expected primary payer for the cohort was Medicare (69.85%).

**Table 1:** Study Demographics.

| **Demographic** | **Cohort Makeup (%)** |
| --- | --- |
| Male | 63.68 |
| Female | 36.32 |
| White | 77.03 |
| Black | 10.6 |
| Hispanic | 7.09 |
| Asian/Pacific Islander | 2.13 |
| Native American | 0.52 |
| Other | 2.63 |
| **Total Patients Studied** | 2,011,854 |
| **Patients with Coronary Perforation** | 11,725 |

Based on univariate analyses of the occurrence of coronary perforation in PCI patients with an age above 30 years old, individuals who experienced coronary perforation had a significantly higher death rate (12.97%) than individuals who did not (2.57%) (Odds Ratio [OR], 5.64; CI, 5.00–6.37; p<0.001). (*Figure 2*). In addition, as perforation rate has increased from 0.11% in 2016 to 0.12% in 2020, perforation death has also increased from 10.76% in 2016 to 13.26% in 2020. (*Figure 1,2*).

**Figure 1.**
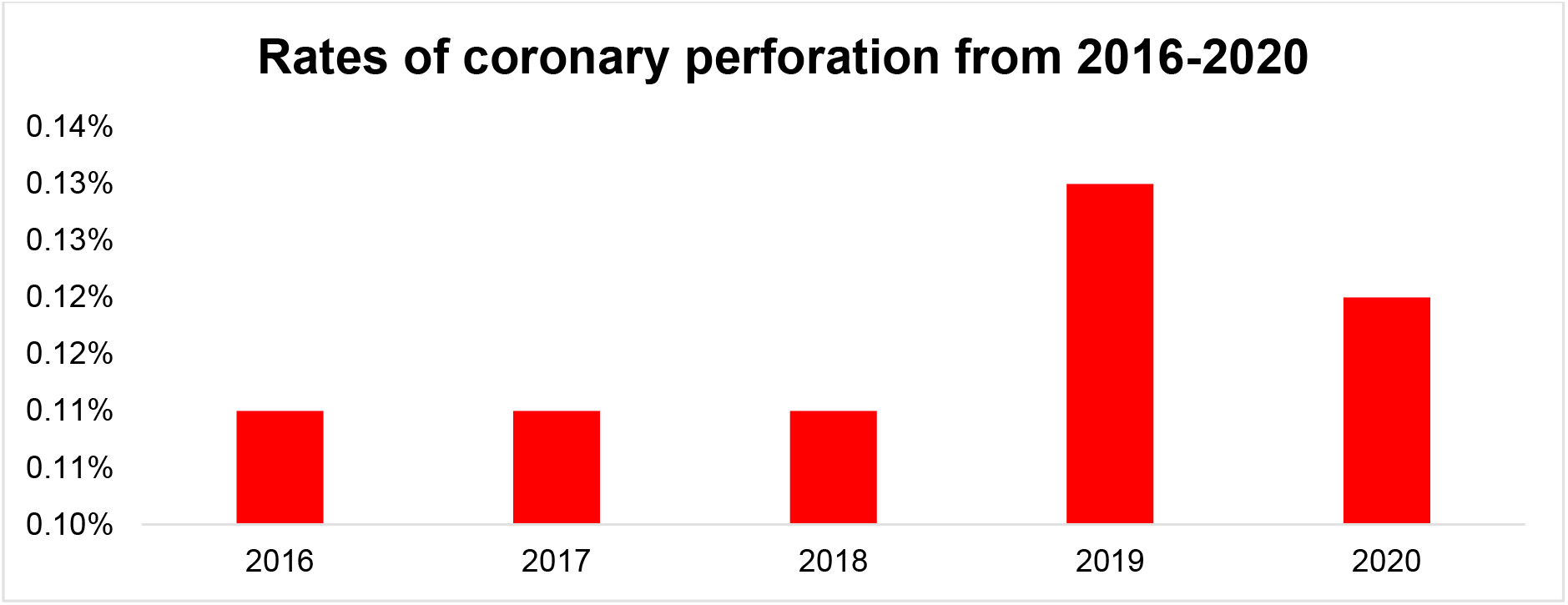
Perforation rates have consistently been 0.11% and above from 2019-2020.

**Figure 2.**
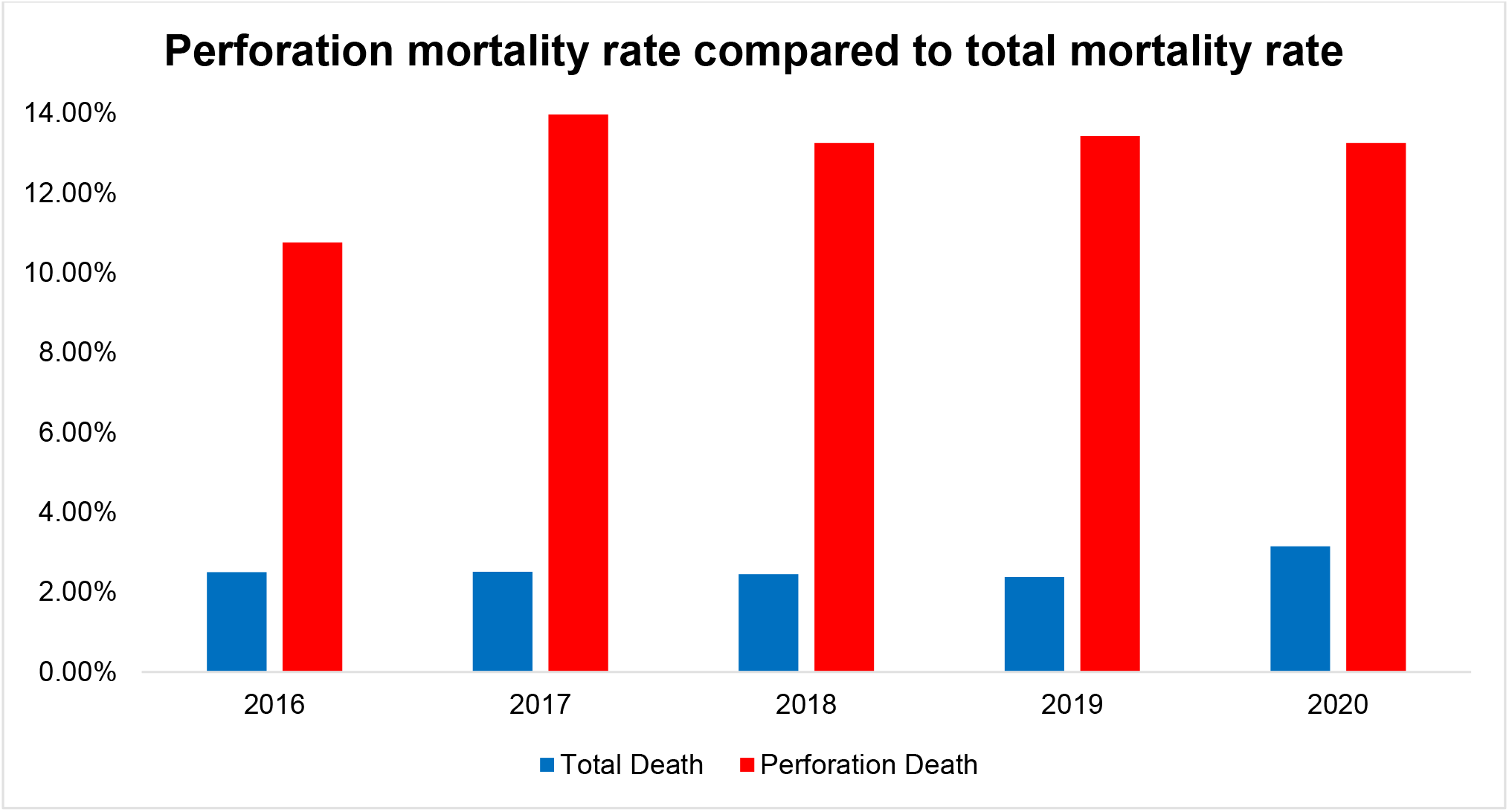
The mortality rate of patients with perforations was very high in comparison to patients without perforations. (12.9% vs 2.5%, OR: 5.6, CI:5-6.3 p<0.001).

In terms of complications, when compared with individuals who did not have a coronary perforation, those that had a coronary perforation as a result of PCI also experienced significantly greater percentages of pericardial effusion (21.19%; p<0.001), cardiac tamponade (21.02%; p<0.001), acute lactic acidosis (12.71%; p<0.001), nontraumatic ischemic infarction of the muscle (0.09%; p<0.001), atherosclerosis of extremity arteries with rest pain (0.72%; p<0.001), postprocedural acute kidney failure (0.43%; p<0.001), acute posthemorrhagic anemia or anemia following acute blood loss (26.14%; p<0.001), acute postprocedural respiratory failure (2.47%; p<0.001), disseminated intravascular coagulation (0.94%; p<0.001), and cardiac arrest (5.37%; p<0.001). The only complication that had a higher percentage of occurrence in individuals who did not have a coronary perforation because of PCI was sepsis (7.02% of individuals without coronary perforation; p<.001). (*Figure 3*).

**Figure 3.**
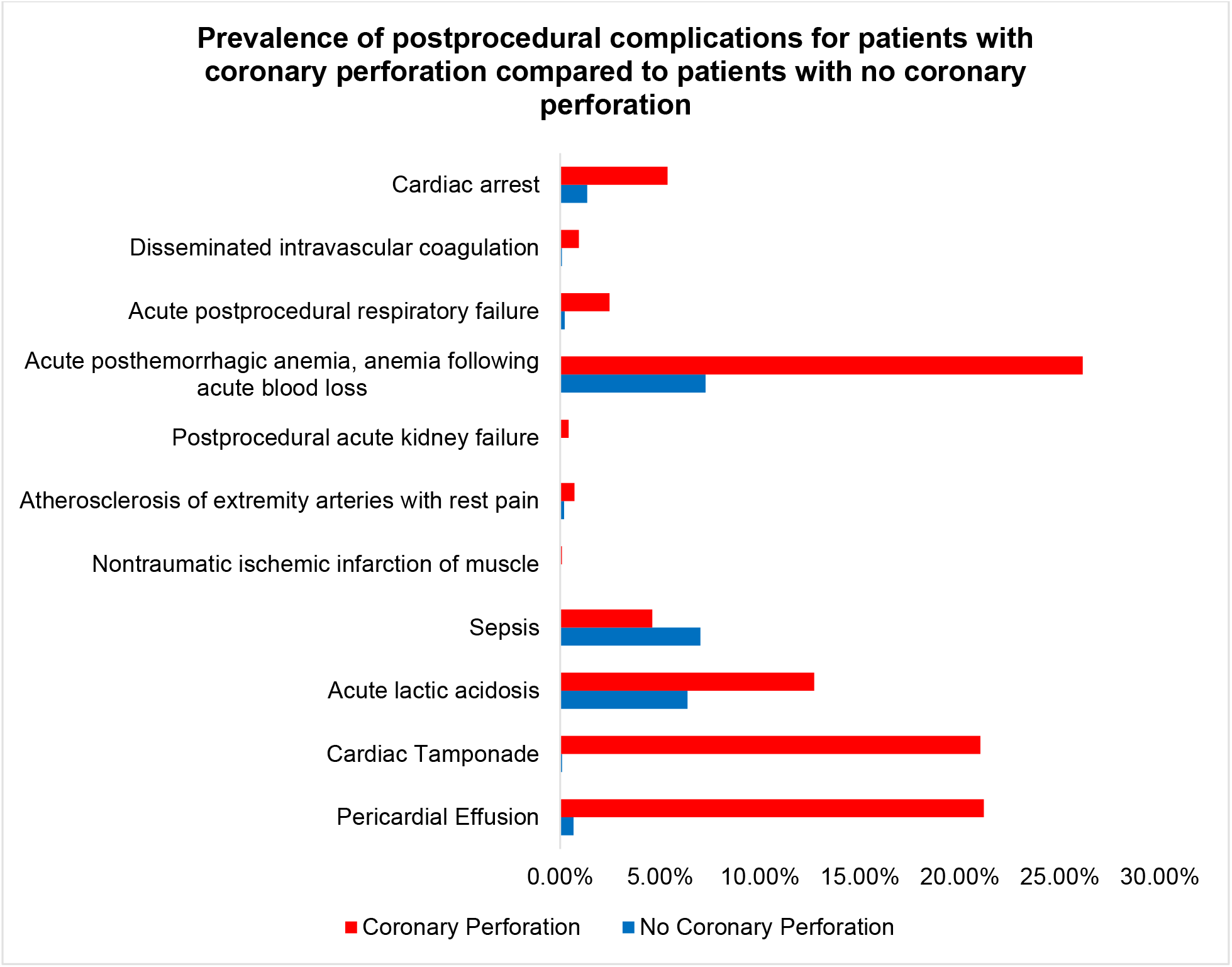
Most post-procedural complications occurred at higher percentages except for sepsis, in patients with coronary perforation than those without coronary perforation.

## Discussion

This study examines the occurrence of coronary perforation and perforation-related mortality and complications in a cohort of adults aged 30 and older from a large inpatients database in 2016-2020. Coronary perforation occurred in 0.12% of patients who underwent PCI. However, despite this low percentage, individuals who experienced a coronary perforation had a significantly higher death rate (12.97%) than individuals who did not (2.57%) (Odds Ratio [OR], 5.64; CI, –6.37; p<0.001). Furthermore, the rate of perforation rate has been steadily over the 4 years study 0.11% in 2016 to 0.12% in 2020) without any reduction in the high mortality rates of over 10% over the years (10.76% in 2016 and 13.26% in 2020) suggesting lack of progress in prevention and treatment of coronary perforations over the years. Current preventative measures for coronary perforations include being careful with wire manipulation during PCI as well as avoiding ballon and stent oversizing ^18^. Restricting the indication of high-risk PCI to those who benefit the most may contribute to a reduction in the perforation rates in the future particularly in patients with chronic total occlusion (CTO)^19^.

Treatment options for coronary perforations include the use of an immediate blocking balloon to prevent extravasation near the site of the PCI at low pressure for 5-10 minutes^19^. Following this, other recommendations include stopping glycoprotein IIb/IIa inhibitors and replacing heparin with protamine ^20^. In addition, more advanced perforations may indicate the need for covered stents to and coil embolization to prevent further leakage of blood and sealing of the site^20,21^.

Despite the known treatment options when a coronary perforation occurs, mortality and complications remain high based on our data suggesting that we need to improve our approach to coronary perforation and treatment in order to reduce persistent high mortality seen in our database over the years.

## Conclusion

Using a large national inpatient database, all-cause inpatient mortality in patients with coronary perforation was found to be very high (over 10%) with persistently high mortality and complications over the year studied. Our data suggest that prevention and treatment of coronary perforations have not improved over the years. Therefore, any effort to prevent perforation with better treatment for perforations can substantially reduce mortality in patients undergoing PCI.

## Limitation

This study has some significant limitations, specifically that we could not separate the coronary perforations by the type of perforation. Having mortality and adverse cardiac event percentages specific to each of the three types of perforation would have led us to make a more precise conclusion on the relationship between PCI-related perforation category and mortality/adverse events. In addition, the study only uses data from open-source reported PCI-related perforations. This could underestimate the true occurrence of perforations and associated adverse events and mortality. Furthermore, we used ICD-10 coding with its inherent limitations.

## Data Availability

NIS database publically available

